# Predicting Common Pathway Signatures Between DNA Methylation and Post Translational Modification in Type II Diabetes & Parkinson’s Disease Using Heterogeneous Data Integration

**DOI:** 10.1101/2024.09.26.24314438

**Authors:** First Saikat Biswas, Second Pabitra Mitra, Third Krothapalli Sreenivasa Rao

## Abstract

The complex diseases, namely, Type 2 Diabetes Mellitus (T2DM) and Parkinson’s Disease (PD), are extensively studied due to their prevalence in a large population group. Between these two diseases, T2DM is denoted as the zero index disease in a patient, which may lead to PD in a more advanced clinical stage. Both of these diseases may occur due to abrupt DNA methylation of genes. Likewise, both diseases may occur in a patient due to protein misfolding. Our study proposes a novel framework for building two disease-specific heterogeneous networks by integrating different tissue-based transcriptomics, epigenetics, epistasis, and PPI-based topological information. We predict the missing links between the DNA methylation and Post-Translational Modification (PTMs) associated with protein aggregation. Next, we have predicted the common signature of the prevalence of linked patterns in both diseases, further validated by relevant biological evidence.

## 1 Introduction

Multi-omics data provides profound insights into the complex connections of different biological mechanisms. These new insights have helped to unravel the hidden causes of complex diseases and their corresponding inter-relationships. In some of the recent works, this multi-omics integration is used to identify biomarkers or pathway features associated with complex diseases [36, 40, 41]. Currently, multi-omics methods are being applied to comorbid diseases like Type 2 diabetes mellitus (T2D) and Parkinson’s disease (PD).

T2D is an increasing health hazard that is closely linked to the epidemic of obesity [16]. The core deficiencies of T2D are insulin resistance and impaired insulin secretion. Likewise, PD is another chronic, progressive neuro-degenerative disease characterized by both motor and non-motor features [17]. Due to its gradual degenerative effects on mobility and muscle control, PD has a significant clinical impact on patients, families, and caregivers [17]. T2D and PD are both comorbid diseases, with T2D as the primary disease. In population-based cohort studies, it is observed that patients with T2D appear to be at high risk of developing PD [13]. A progression and a more severe phenotype of PD share common cellular pathways [11, 19]. A multi-omics-based approach is currently being followed to find out the increased fasting plasma glucose (FPG), level as a potential biomarker in T2D [24]. Another work explains the functional alterations of the liver in insulin-deficient T2D patients using the multi-omics technique [5]. Similarly, the effectiveness of multi-omics is noticed in PD [32, 62]. Redenšek et al. [62] in their recent work has analyzed the hidden molecular mechanisms of PD using thirteen omics layer data. Similarly, biomarker identification of early-onset PD prognosis is performed using a multi-omics joint analysis approach [32]. Epigenetic variations add a new perspective to the above studies [69].

Integration of gene expression with DNA methylation refers to the combination of transcriptomics and epigenomics level data. This kind of unification of two omics data has well served the purpose of efficiently retrieving the hidden biological features for complex diseases [44, 69, 72]. DNA methylation is one of the most elementary aspects of this epigenetic variational study, due to its proven influence in a variety of human complex diseases [37, 47, 48]. The reason behind DNA methylation’s claim to be the most characterized epigenetic modification is its influence on gene expression via the disruption of transcription factor (TF) binding in the promoter regions and the recruitment of methyl binding proteins, that introduce chromatin compaction along with gene silencing [69, 83]. Aberrant methylation can affect the functions of tumor suppressor genes by altering their expression levels in CpG islands located near the promoter region of the genome. Recently, [2, 87] analyzed the role of DNA methylation in the pathogenesis of T2D in humans. Miranda et al. [55] have extensively studied the implications of DNA methylation in PD. In their work, they have shown that DNA methylation takes a crucial role in gene-environment interactions associated with PD. Other very recent works regarding the participation of DNA methylation in PD are also well-studied [56, 58].

Like DNA methylation, protein misfolding is another important condition that causes several complex diseases including diverse systematic disorders T2D, and neurodegenerative disorder PD. Misfolding of protein, which is either degraded or aggregated in the cell, leads to the causation of diseases like T2D and PD [26, 68].

Heterogeneous network represents a large network embodying multiple information. Here the nodes represent different types of entities and their corresponding edges stand for distinct relationship types. This heterogeneous information can range from genomics proximity to co-expression, from experimentally validated topological interactions to associations mined from medical literature. The advantage of using a heterogeneous network in systems biology is to logically infer the hidden patterns or underlying connections within it from the collective information. In specific, the integration of data sources into a heterogeneous network and the logical application of appropriate search algorithms lead to the discovery of hidden novel links, and yet unknown patterns [65]. In recent work, Tran et al. [73] have predicted various novel disease-gene associations using a heterogeneous network with node kernels. Another heterogeneous network-based approach is suggested by Sügis et al. [70] to predict the hidden disease mechanisms of Alzheimer’s disease. Our proposed heterogeneous network for comorbid disease pattern analysis is constructed by integrating gene expression, pathway, DNA methylation, single nucleotide polymorphism, amyloid precursor proteins, and post-translational modifications.

## 2 Related Works

We review a few studies that showed the analysis of T2D and PD from DNA methylation and protein aggregation perspectives. Recently, a work by Kim [43] has suggested that cumulated errors in DNA methylation lead to altered gene expression, which affects the response to external stimuli and causes the development of T2D [23]. According to this work, passenger DNA methylation is crucial for T2D progression. Another work shows the methylation of CpG sites in regulatory regions outside the gene promoter region also plays a critical role in regulatory tissue-specific gene expression [2]. One of these regulatory elements is the cycle adenosine monophosphate (CAMP) responsive element (CRE), which can bind with a diverse array of transcription factors [28]. This proposed work shows that the methylation of CpG sites within CRE reports to independently suppress insulin promoter activity by approximately 50%. Similarly, the effect of DNA methylation on causing PD has recently been observed in a few works [29, 56, 79]. These studies show that methylation of *αsynuclein (SNCA)* and microtubule-associated protein *tau* gene appear to be of special interest and DNA methylation effect is also observed in neighboring other candidate genes responsible for causing PD. One of these works performs cross-sectional analysis by comparing methylation profiles between cases and controls. Their method has identified differentially methylated position(s) (DMP) in PD samples with significant P-values (P *<* 5.0E-7). Another important cause of T2D and PD is protein aggregation or protein misfolding, which is also extensively studied in a few current works [26, 56, 57]. Chiti et al. [12] have suggested that amyloid deposits in the brain of PD patients contain 140-residue *α-synuclein (SNCA)*, and T2D pancreatic amyloid deposits contain total thirty-seven residue islet amyloid polypeptides (IAPP). The predominant evidence suggests that native non-toxic monomers of *α-synuclein (SNCA)* in PD and human islet amyloid polypeptide (hIAPP) in T2D, may misfold and self-aggregate into *β*− *sheet* enriched soluble cytotoxic oligomers. These oligomers further elongate into pathogenic insoluble fibrillar assemblies [18], and the condition leads to these complex diseases. In recent work, Ahmed et al. [3] have shown that defective protein folding and DNA methylation in CpG sites cause T2D. According to their claim, the basis for this disease is attributed to oxidative stress, chronic inflammation, non-enzymatic glycation of proteins, and epigenetic changes.

Heterogeneous multi-modal approach on different biological information infers the novel hidden disease mechanisms of Alzheimer’s disease in the work suggested by Sügis et al. [70].

In numerous current link prediction tasks, the graph convolutional neural network (GCN)-based semi-supervised classification technique by Kipf et al. [45] has demonstrated outstanding success. The obtained results from this prediction have shown high accuracy and are executed with good speed for considerably large graphs.

We propose a new way to predict and study the hidden patterns of comorbidity between two common diseases by combining different types of biological data into a single heterogeneous network. This network will be used for both T2D and PD. As the combined information from transcriptomics, epigenetics, and epistasis can help to understand better the proper biological and pathological inter-relations between different biological entities, we propose a unique approach to build entity-type specific subnetworks with different tissue-oriented genomic information and combine them to infer the common target pattern from this aggregated information using heterogeneous graph neural network. Thus, the novel contributions of our work are:

1. We integrate the different tissue-specific omics data for T2D and PD to build individual complex heterogeneous networks.
2. We predict the connecting pathway signatures between T2D and PD based on these heterogeneous networks regarding DNA methylated genes and post-translational modifications.

## 3 Materials and Methods

This section describes the data and the details of the proposed approach.

### 3.1 Data description

A heterogeneous biological network is a multi-model approach, where a network is formed by integrating the individual datatypes to infer a systematic view of disease.

In this work, we build heterogeneous networks individually for T2D and PD. We combined the expression-level information and the topological data to build the network. In particular, the heterogeneous network is made with the microarray gene expression datasets for T2D (GSE64998 [46]) and PD (GSE99039 [64]). The tissue samples used for gene expression profiling for T2D and PD are liver and whole blood samples corresponding to the control and treated groups. The microarray-based DNA methylation profiling used for T2D is GSE65057 [46] and GSE111629 [14, 15, 30] for PD. The tissue samples, namely, liver and whole blood, are used for methylation profiling for T2D and PD, respectively. The Illumina Infinium 450k Human DNA methylation BeadChip is used for both of the diseases. Similarly, the genes from the corresponding proteins are obtained from the UniProtKB database. The other different biological databases used for the construction of heterogeneous networks are defined below :

- Human biological signalling pathway interactions are obtained from MSigDB c2 (canonical pathway) database [52], downloaded on 18th July, 2020. We get a total of 2200 sets of biological metabolism and signaling pathways.
- The central *invivo* and *invitro* seed proteins for building amyloid interactome are taken from the work proposed by Biza et al. [8].
- The human protein-protein interaction (PPI) is retrieved from IntAct (03-2020) in MITAB 2.5 format file [39]. We obtained a total of 1,60408 protein-protein interactions from the final parsed file.
- The associations between protein and their corresponding post-translational modification (PTM) interactions are obtained mostly from dbPTM database [34, 51] and BioGrid [10] and a few from UniProtKB. Specifically, the protein-PTM associations for the PTMs, namely, acetylation, amidation, hydroxylation, methylation, N-linked glycosylation, phosphorylation, S-nitrosylation, sumoylation, and ubiquitination, are obtained from dbPTM and BioGrid databases. The remaining PTMs, namely, proteolytic cleavage, carboxylation, disulfide bonds, and oxidation-related protein associations, are extracted from the UniProtKB database.
- The single nucleotide polymorphisms (SNPs) for the mapped traits, namely, T2D and PD, are downloaded from the NHGRI-EBI catalog of human genome-wide association (GWAS) studies [All associations v1.0.2] [9].
- Associations for PTMs with the corresponding SNPs are obtained from AWESOME database [81].

The complete heterogeneous network proposed in this work is shown in the supplementary Figure S1. Next, we describe in detail the individual data components. The schema of steps for data processing, building individual sub-networks, and integrating them to build a heterogeneous network (for each of the diseases) to predict common hidden patterns is shown in Fig. 1.

**Fig. 1:**
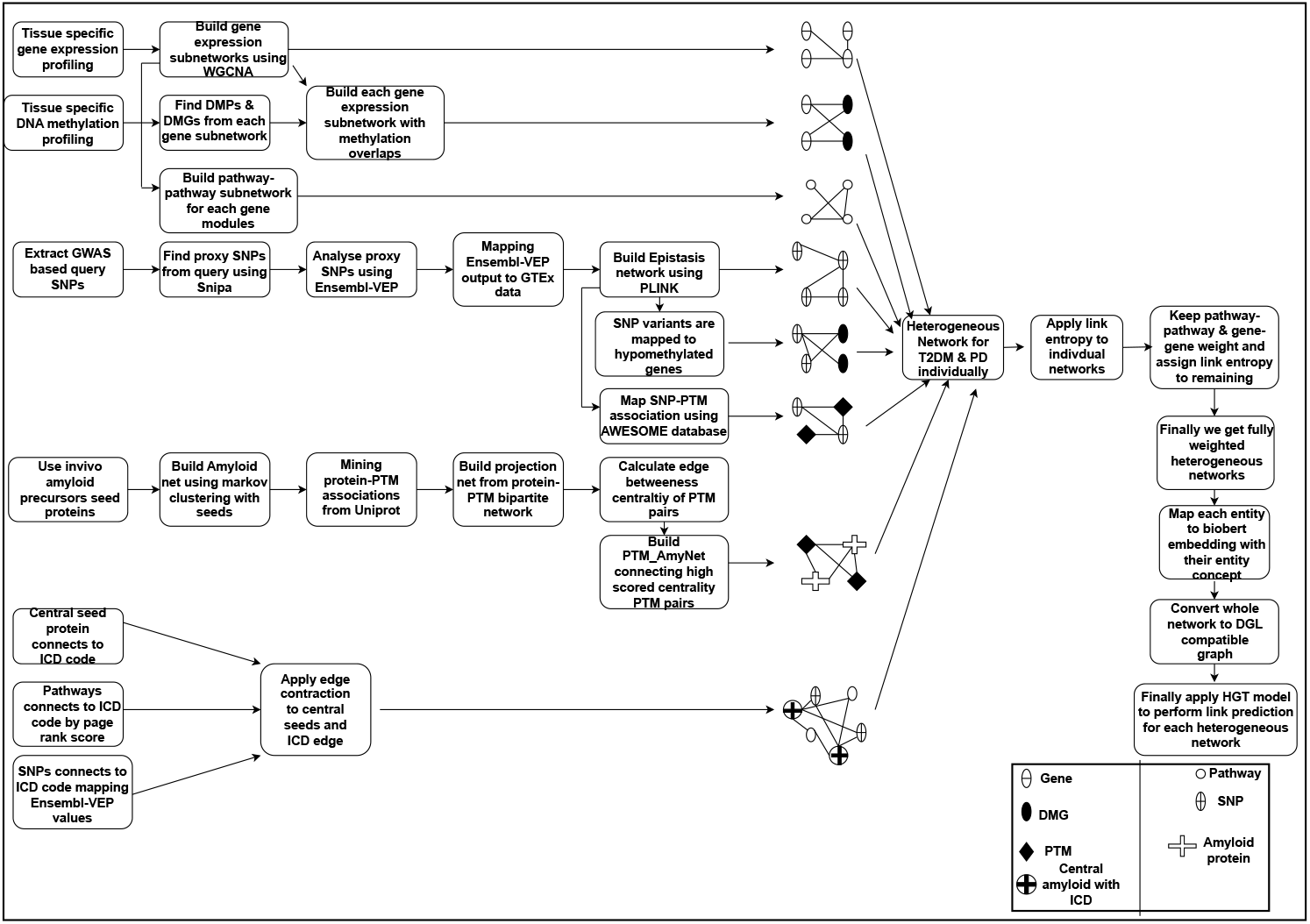
The workflow of link analysis between DNA methylation and post-translational modification

### 3.2 Gene expression profiling

Gene expression profiling is a method to determine the pattern of genes expressed. This gene expression is studied at the transcription level, under conditioned specific circumstances, or in a specific desired cell to get a global picture of cellular functions for further molecular-level analysis. In our proposed work, DNA microarrays are used individually for T2D and PD. To analyze the profiles, R packages namely, oligo, oligoclasses, GEOquery, limma, hugene11sttranscriptclusters.db, pd.hugene.1.1, Org.HS.eg.db, annotate are used. The CEL file and the corresponding series matrix file are loaded. The steps for expression profiling for T2D and PD are defined below:

- The phenodata of both of the diseases are retrieved from series matrix files and CEL files for 21 samples (GSE64998) for T2D and 558 samples (GSE99039) for PD are read.
- The obtained phenodata for T2D gives information of metabolically healthy, obese non-diabetic, and obese type II diabetic patient samples. The phenodata regarding PD gives information on healthy controls, Idiopathic Parkinson’s disease, genetically unaffected controls, Parkinson’s disease with a genetic cause, etc.
- The phenodata and the CEL files individually for both of the diseases are mapped together.
- Both of the microarray data are normalized using rma (robust multiarray average) based method.
- To get the differentially expressed genes (DEGs) for T2D and PD, the normalized expression sets are used to build the design matrix for them and the differences of control and disease states from them are used to construct the contrast matrix.
- The obtained contrast matrices are then fitted in a linear model.
- Finally, the obtained DEGs are sorted accordingly with their logFC values in ascending order and corrected P-value of *<* 0.05.

### 3.3 Gene co-expression modules using Weighted Gene Co-expression Network Analysis (WGCNA)

To find out the correlation pattern among genes across microarray samples, gene-correlation network analysis has already shown a greater impact [84]. On this matter, Langfelder et al. [49] have proposed a weighted correlation network analysis method for finding clusters (modules) of highly correlated genes. Their technique is widely used in finding out important clusters using the module eigen genes or intra-modular hub genes.

To build the proposed heterogeneous network, *WGCNA*[49] R package is used. WGCNA builds the topologically correlated gene expression modules from the normalized gene expression set dataframe. The functions used for WGCNA analysis regarding the proposed work can be divided into (a) network construction and (b) individual module detection.

Each of these steps is explained as followings:

#### 3.3.1 Network construction

A network is well specified by an adjacency matrix *a*_*ij*_, a symmetric *n × n* matrix with entries [0,1], whose individual component *a*_*ij*_ encodes the network connection strength between nodes i and j. An intermediate quantity, called co-expression similarity *S*_*ij*_ is defined to calculate this adjacency matrix. The function adjacency calculates the adjacency matrix from our filtered expression data of T2D and PD.

#### 3.3.2 Individual module detection

After the network is constructed, the topologically correlated modules are detected. Here, the modules refer to the densely interconnected gene clusters. The module detection operation is done by the following steps:

- First, the topological similarity is calculated from the adjacency matrix.
- Next, hierarchical clustering is used on the obtained result in the previous step.
- The modules of biologically related correlated genes are obtained.
- The module-specific eigen genes are identified using the *moduleEigengenes* function.
- The correlation is again calculated using these obtained module eigen genes and the hierarchical clustering is applied again on the previously calculated dissimilarity of the correlation.

The aforementioned steps are applied for both diseases, T2D, and PD. After applying this WGCNA method finally we get 57 and 21 gene-coexpression modules for T2D and PD respectively.

### 3.4 Differential methylation profiling

The recent advances in transcriptomics data have helped to focus on the statistical analysis of DNA methylation data for a deep understanding of epigenomic activities in a cell. In our proposed method we have used GSE65057 and GSE111629 microarray-based methylation data for T2D and PD respectively. The analysis is performed using R packages namely, *GEOquery, minfi, limma, IlluminaHumanMethylation450kanno*.*ilmn12*.*hg19, IlluminaHumanMethylation450kmanifest, missMethyl*, and *DMRcate*. The steps for performing DNA methylation experiment are as below:

- At the first step, the methylation data is used to build the expression set, and the metadata is extracted from the series matrix using the GEOquery package.
- The extracted metadata is then mapped to the expression set to the sample-specific group status (i.e. control and disease) and associated tissue information.
- The expression set is then sorted according to their p-values *<* 0.05.
- The samples with poor quality are discarded and the remaining sample expression set is extracted.
- The expression set matrix is normalized using *processQuantile()* function as in the methylation profile only one type of tissue namely, the liver in T2D and whole blood are used for PD.
- In the next step, the M-values, and *β*-values are calculated from the normalized expression set. The M stands for methylation intensity and *β* refers to the ratio of methylation intensity and summation of methylation and unmethylation intensities.
- The M-values and *β*-values are calculated using *getM()* and *getBeta()* functions respectively.
- Now the design matrix is formed using the control and disease status. For, T2D these different labels are obese non-diabetics, obese type II diabetics, and metabolically healthy individuals. Whereas, for PD these labels are PD-free control and Parkinson’s disease associated with female and male patients.
- Finally, the differentially methylated positions (DMPs) are obtained for the desired coefficient with their corresponding M-values and sorted accordingly with their P-values.
- Next, we extract the differentially methylated regions (DMRs) using *DMRcate()* package by fixing the parameters lambda = 1000 (lambda denotes the gap value between significant CpG sites) and scaling factor for bandwidth fixed to 2.
- To get the multiple comparisons Benjamini-Hochberg (BH) [7] based FDR (False Discovery Rate) is used, where values below 0.05 are considered significant.

Now, we extract the overlapping DMPs and DMRs with the genes in the gene-coexpression modules obtained using the WGCNA method individually for T2D and PD.

### 3.5 Pathway sub-networks on the basis of individual gene-coexpression network modules

The pathway sub-network building for individual gene-coexpression modules is inspired by the concept proposed by Zheng et al. [86]. The whole method of sub-network formation is divided into the following explained steps.

#### 3.5.1 Projecting the candidate genes from each gene-coexpression network modules to individual pathways

In our proposed work, the candidate genes from all 57 gene-coexpression modules for T2D and 21 gene-coexpression modules for PD are extracted individually to get the module-specific collective genes. The biological signaling pathways are obtained from the database MSigDB C2 (canonical pathway), from which we obtain a total of 2199 sets of biological metabolism and signaling pathways. The module-specific candidate genes are projected into the pathways to calculate the activity score of each pathway.

The gene expression values are retrieved from the normalized gene expression value set obtained earlier for each of the candidate genes collected from the corresponding unique coexpression modules. The activity scores are calculated according to these expression values of the candidate gene set on each sample. The gene expression value of a gene in the biological pathway for every sample constitutes an expression value matrix; where each column represents a sample and each row represents a gene. A biological pathway includes *N*_*p*_ genes, whose expression values can form a matrix of *N*_*p*_ rows. Each biological pathway refers to an activity vector, and the dimension of the vector is the number of samples, that is in each sample *j*, an activity score can be calculated. The calculation formula is given below:

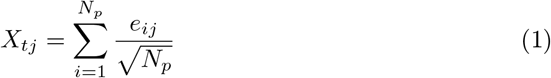

Where, *X*_*tj*_ represents the activity score of the *t* biological pathway in the *j* sample, and *e*_*ij*_ represents the gene expression value of the *i* gene in the *t* biological pathway of the sample *j*, and *N*_*p*_ is the number of genes in the biological pathways; vector [*X*_*t*1_,*X*_*t*2_,…, *X*_*tm*_] represent the activity score of the *t* biological pathway in m samples. Then a phenotype vector is formed based on the phenotype labels of the samples, and then the mutual information between the activity vector and the phenotypic vector of the samples (control and case) is computed by combining the activity vectors of each biological pathway.

#### 3.5.2 Mutual information with the phenotype

Mutual information (MI) is the most commonly used method of information theory, meant for information calculation. By calculating the MI between activity vectors and phenotypic vectors of the respective biological pathways, the correlations between the two vectors are measured. This shows the influence of a biological pathway on the phenotype of T2D as well as the same method applied to PD. Constructing a phenotype of the sample [*C*_1_,*C*_2_,…, *C*_*m*_], the phenotype vector is a zero-one vector, where if the sample is diabetic it must be 1 otherwise it is 0 and a similar convention is also followed for PD and non-PD samples as well.

Using *X*(*i*) to indicate the activity score of the *i*^th^ biological pathway to each sample, *X*(*i*) = [*X*_*i*1_,*X*_*i*2_,…, *X*_*im*_]. C is used to define the phenotype vector of m samples, C = [*C*_1_,*C*_2_,…, *C*_*m*_]. Thus, the correlation between the biological pathways and disease phenotypes S(i) can be represented by mutual information, MI(*X*^*′*^ (*i*),*C*) between *X*^*′*^ (*i*) and *C* vectors. Here, *X*^*′*^ is the discretized form of *X*. The applied formula is given below:

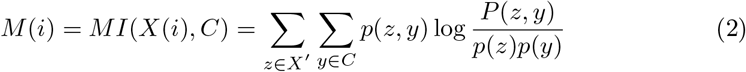

Where, the activity score *X* is discretized into median value based equally spaced bins to obtain the *X*^*′*^, respectively, *p*(*z, y*) is the joint probability density function of *X*^*′*^ and *C* and *p*(*z*) and *p*(*y*) are the marginal probability density function of *X*^*′*^ and *C*. The whole method is applied individually for all of the gene-coexpression modules.

#### 3.5.3 Gene-coexpression module specific pathway network construction

The module-specific pathway-pathway interaction can be represented as a module-specific sub-network, which is constructed as follows:

Let, the subgraph *H*_*mk*_(*V*_*mk*_, *E*_*mk*_) is comprised of a set *V*_*mk*_ of module specific pathways and a set *E*_*mk*_ denotes the module specific weighted pathway-pathway interaction network with *E*_*mk*_∈’ *V*_*mk*_ ∗ *V*_*mk*_. To define this weight the logarithm of the summation of phenotypic mutual information (calculated previous section) and semantic similarity is taken. Here, *V*_*mk*_ = {*P*_*mk*1_, *P*_*mk*2_,…, *P*_*mkn*_} and n = |*V*_*mk*_|. Matrix A represents the weighted *n× n* adjacency matrix of *H*_*mk*_; where k is the number of modules (gene-coexpression modules). For an instance say, two pathways *P*_*mk*1_ and *P*_*mk*2_ are the set of genes from the module *m*_*k*_ participating in the corresponding pathways.

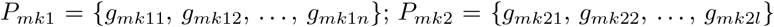

The semantic similarity is measured by the mutual sharing of GO (Gene Ontology) functions among the module-specific pathway pairs. The calculation is done by the following :

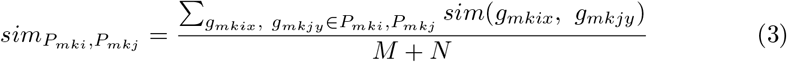

Where, M = |*P*_*mki*_| is the number of genes on the pathway *P*_*mki*_ and N = |*P*_*mkj*_| is the number of genes on the pathway *P*_*mkj*_.

We have used the *GoSemSim* R package for the semantic similarity calculations. The semantic similarity *sim(g*_*mkix*_,*g*_*mkjy*_) between genes, refers to the similarity on the basis of Molecular function (MF), Biological process (BP), and, Cellular component (CC). Wang’s method [75] is used to calculate the semantic similarity in the proposed work. The edge weight calculation is performed for all the pathway pairs of each module of obtained all 57 gene-coexpression modules for T2D and all 21 gene-coexpression modules for PD. To calculate the final edge weight of each module-specific pathway-pathway network, we follow a specific trick to integrate the phenotypic mutual information and semantic similarity scores. This technique calculates the maximum of pairwise pathways and summed up with the corresponding semantic similarity of the pathway-pathway interaction. For instance, say, we have two pathways *p*_1_ and *p*_2_ and their corresponding mutual phenotypic weights are *wp*_1_ and *wp*_2_, respectively. The semantic similarity score between these pathways is *w*_*sem*_. Then the mutual phenotypic information weight is calculated as the following:

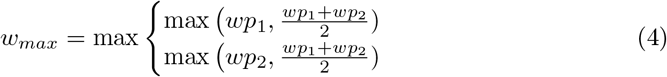

So, the final weight of the corresponding edge between *p*_1_ and *p*_2_ is :

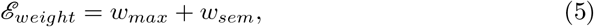

where, ℰ _*weight*_ is the interaction weight between the pathways, *p*_1_ and *p*_2_. This pathway-pathway edge weight calculation is also applied similarly to PD.

### 3.6 Amyloid protein-protein interactome construction using seed proteins

To build the amyloid protein-protein interaction (PPI) network, we first parse the PPI data from IntAct (05-2016) is retrieved in a MITAB 2.5 format file [39] with two columns of gene names. Then, we apply the Markov Clustering (MCL) algorithm [20] to obtain the clusters of proteins. We get the 28 invivo amyloid proteins, the most critical central proteins responsible for amyloid fibril formation. The work by Biza et al., [8], is the source of all 28 seed amyloid precursor proteins. These invivo amyloid proteins are then mapped to their corresponding UniProtKB gene names. We then search for the clusters (obtained using MCL), which contain at least one or more than one seed protein. The clusters containing seed protein(s) are combined to get the final desired proteins to build the amyloid interaction network. These proteins are then searched through the IntAct database for related interactions. We group all these PPIs to get the final amyloid protein interactome. We finally get a total of 46,634 interactions. We refer to this network as AmyNet.

### 3.7 Mapping of PTMs with corresponding proteins and building PTM-AmyNet

Post-translational modifications (PTM) are essential in different stages of amyloid formation. It is already a known fact that enzymatic PTMs are extensively linked to protein misfolding or protein aggregation and cause amyloid dispositions in pathological conditions. When enzymatic PTMs result in excessive or differential modifications, they can adversely impact the propensity of protein and lead to protein aggregation. So PTM and corresponding protein associations are important, as their collective information can shed light based on PTM participation in protein misfolding.

In this method, we have used 14 types of PTM associations. These are (i) Acetylation, (ii) Amidation, (iii) Hydroxylation, (iv) Methylation, (v) N-linked Glycosylation, (vi) O-linked Glycosylation, (vii) Phosphorylation, (viii) S-Nitrosylation, (ix) Sumoylation, (x) Ubiquitination, (xi) Carboxylation, (xii) Disulfide-Bond, (xiii) Oxidation, and (xiv) Proteolytic Cleavage. To obtain the PTM corresponding protein associations, we search through the dbptm database. From dbptm, we get the protein associations to the PTMS, namely Acetylation, Amidation, Hydroxylation, Methylation, N-linked Glycosylation, O-linked Glycosylation, Phosphorylation, S-Nitrosylation, Sumoylation and Ubiquitination. But for the Proteolytic Cleavage, Disulfide Bond, Carboxylation, and oxidation we mined the protein associations from the UniprotKB database. Next, we search all these PTMs PTM-associated proteins in our AmyNet network to obtain those proteins’ corresponding interactions. Finally, we build the PTM-AmyNet (PTM-Amyloid protein Network) network by integrating the PTM-protein associations with the AmyNet interactome.

#### 3.7.1 Building PTM projection network from the bipartite network with PTMs-Protein associations

The bipartite network from PTMs to protein associations is built using the python *Networkx* package. The aim of building this bipartite network is to obtain the PTM projection network from it. After getting the PTM projection network, we calculate the edge-betweenness centrality of the PTM-PTM pairs. We used the algorithm proposed by Girvan et al. [25] to obtain the edge betweenness centrality scores. These scores are sorted in descending order, and only the highest centrality PTM pairs are taken. For instance, if we get more than one PTM pair with similarly high scores, all the same high-scoring pairs or edges are considered. Finally, the PTM_AmyNet network connects the PTM pairs with these high edge betweenness centrality scores.

### 3.8 SNP variant analysis and building SNP-SNP epistasis network

The corresponding query SNPs are extracted and parsed to get the GWAS-based query SNPs for mapping traits as T2D and PD. These obtained SNP variants are then used to collect the proxy SNPs by using snipa https://snipa.helmholtz-muenchen.de/snipa3/ [4] online tool. The analysis is being performed by using the LD≥ 0.8 (linkage disequilibrium). Thus, we get the desired proxy SNPs with their corresponding minor allele frequencies. The consequence types, alternative, and reference allele information with their corresponding phenotypic and disease associations of the acquired proxy SNP variants are obtained using Ensembl-VEP [53] tool. As our performed experiments are based on the liver tissue and whole blood tissue for T2D and PD, respectively, of the European population, we now matched the VEP SNP outputs with the GTEx (Genotype-Tissue Expression) [1] European population-based dataset. Finally, we obtain the GWAS-based SNP variants for tissue-specific European population-based Ensembl-VEP results for each of the diseases. From these SNP variants with corresponding minor allele frequency (MAF) ≥ 0.05 are only selected for analysis, and the remaining are discarded.

The reason to select MAF≥ 0.05 is to find the actual SNP variants that affect population genetics by causing complex diseases. To get the epistasis interaction (SNP-SNP interaction), the PLINK [60] analysis tool is used. The PLINK-based analysis always requires only PLINK-associated genotypic data formats and results to perform its analysis. The corresponding PLINK analysis is done for case-control analysis for the proposed work. These associated data formats are .ped/map extended file formats for the corresponding case-control-based genotypic and phenotypic information. These specific types of file creation further need the base files of three particular results, which are in .bim, .fam, and .bed file formats. Researcher Alejandro Ochoa recently built an R package, namely, *genio* (Genetics Input/Output Functions) [82] to analyze and create allele-specific population genetic data for SNP variants. In this analysis, we take the same number of cases as in the expression set along with the randomly shuffled 20%

### 3.9 Extracting Hypo/Hyper methylated genes and connect to SNPs

The differential methylation profiling obtained from subsection 3.4 gives the profiling results for T2D (GSE65057) and PD (GSE111629). Individually, after getting the DNA methylated DMPs (differentially methylated positions) and DMRs (differentially methylated regions), the *gometh()* and *goregion()* methods are used from *missMethyl* R package. These methods help to perform the enrichment analysis of the CpG-associated genes. The following steps are performed to extract the hypo/hypermethylated genes:

- We perform the gene enrichment analysis to get the overlapping genes with DMRs.
- Now, the bocals or *β*-values of the CpG-specific data frame obtained earlier are divided into case and control groups by mapping with their metadata information.
- For T2D, the case-control study is being performed between Obese_type2_diabetes and Metabolically_healthy patient samples. Similarly, the case-control distinction is performed for PD based on disease and PD_free_control individuals.
- From each disease-specific CpG data frame, *β*-value row means are calculated, and the absolute difference between the row mean *β*-values of each CpG is measured based on case-control samples. This row mean *β*-value is called Δ *β*.
- We concatenate the CpG specific Δ *β* values with the associated entire dataframe.
- Δ *β >* 0.2, the associated CpGs are hypermethylated, and the CpGs associated with Δ *β <* 0.2, are referred to as hypomethylated.
- We make two class-specific CpG divisions and perform the DMP along with DMR enrichment analysis using *gometh()* and *goregion()* methods, respectively.
- From the obtained results, the CpG entries with P-value *<* 0.05 are considered for further analysis.
- Now, previously obtained genes from each of the gene-coexpression modules, are mapped with these DNA-methylated genes.

In the overlapping of these gene-DNA methylation associations, we get a total of 135 and 169 genes hypomethylated in T2D and PD, respectively. No overlap for each of the diseases is found for hypermethylation.

The SNPs from European population-based liver tissue (for T2D) and whole blood tissue (for PD) related genetic associations are mapped with the integrated SNP variants from the epistasis network. Then, obtained SNP variants are mapped with the acquired hypomethylated genes and the Ensembl-VEP (Ensembl Variant Effect Predictor) [53] generated findings. Finally, these selected SNP variants from the epistasis interaction network are linked with the VEP overlapped DNA methylated genes.

### 3.10 Connecting SNP and PTM nodes using AWESOME dataset

The SNP variants that affect the PTMs are analyzed and predicted by Yang et al. [81] in their AWESOME database. So, the interconnection between the epistasis network and PTM-AmyNet can be initiated by these SNP-PTM interactions. We used this AWESOME dataset to connect these two sub-networks. To get all the SNPs from the epistasis interaction, all the SNPs are integrated and mapped to the parsed result of the AWESOME dataset. The previously mentioned 14 PTMs are only mapped to the data, and the rest have been discarded. Among them, in very few of these interactions, only 22 interrelations of SNP variants and their corresponding PTMs are obtained from these mappings for T2D, and only 6 interrelations of SNP variants are found to be associated with PTMs in PD.

### 3.11 Selective interactions for individual networks with ICD-10-CM codes E11.8 (T2D) and G20 (PD)

The selective interactions with the ICD-CM code E11.8 and G20 for T2D and PD are divided into two kinds of information in the network. One is the global information PTM-AmyNet, and the two local information collectively pathway-pathway interaction and SNP-SNP interaction. As a result, the following three interactions help to identify these interactions:

- **Central Amyloid proteins to ICD -** The invitro and invivo seed amyloid precursors are the central proteins of the PTM-AmyNet network because these critical seeds are used to populate the whole Amyloid protein sub-network. Hence, we only connect these central seed proteins to ICDs E11.8 and G20 for T2D and PD, respectively.
- **Pathways to ICD -** It is a well-known fact that the causation of any disease is highly related to its corresponding pathway relations. So, in the proposed work, these module-specific pathways are also connected to the ICD code.The pathway nodes with the highest page-rank-based centrality scores from each module-specific network are selected.
- **SNPs to ICD -** The Ensembl-VEP output for T2D and PD individually gives the SNP variant information with their DisGeNET-specific known disease associations. Then, the SNPs from the obtained epistasis interaction are integrated to map with these VEP SNPs.

### 3.12 Edge contraction for the sub-networks associated with the ICD-10-CM nodes (T2D:E11.8 and PD:G20)

As in the heterogeneous networks for T2D and PD, there is only one node type, ICD-10-CM-E11.8 and ICD-10-CM-G20, respectively. We combine this single node type with another one using an edge contraction strategy. Edge contraction is used to exclude redundant edges in the network without losing any vital information. To follow this, we select the central amyloid seed proteins for individual networks as the nodes for combining the ICD-10-CM codes. The central amyloid proteins are essential amyloid precursors responsible for protein misfolding in co-morbid diseases. The pathway to ICD entity links and SNP to ICD entity links are connected to this modified node type in each network. In the case of T2D, the ICD-10-CM-E11.8 node type is combined with the 32, and for PD, the ICD-10-CM-G20 node type is combined with 40 different central amyloid precursors. The selected pathway nodes of each module and SNP nodes are connected with these combined nodes.

### 3.13 Integration of all interactions and assigning weights to unweighted edges for individual heterogeneous network

To build the heterogeneous network individually for T2D and PD, all the corresponding subnetworks (i.e., intra-connections) are combined with their associated interconnections.

The subnetworks are, namely, (i) module-specific pathway networks, (ii) gene coexpression module networks, (iii) case-control-based SNP-SNP epistasis interaction networks, and (iv) amyloid PPI along with PTM integrated network. All these subnetworks are linked up with their inter-associated networks. This obtained heterogeneous network is partially weighted because, among all interactions, only pathway-pathway network and module-specific gene coexpression networks are weighted. So, to give weight to the remaining unweighted interactions, a new strategy, namely, link entropy, is introduced in our proposed method. This link entropy method is proposed by Qian et al. [61] in their recently published work. This link weightage technique is divided into two strategies, namely (i) NMF (Non-negative Matrix Factorization) [**?** ], proposed on the whole network topology, and (ii) QS (Quantification Strategy), which is finally used to calculate the LE (Link Entropy) values of all the edges to quantify their significance edge weights. These two strategies are briefly explained in the following sections:

#### 3.13.1 NMF (Non-negative Matrix Factorization)

In this link entropy calculation, we initially consider the whole network unweighted, representing an adjacency matrix A of 1s and 0s. Now, it is assumed that the pairwise interactions in A are influenced by an unobserved expectation network, say, Â. This Â can be defined as:

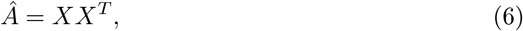

Where the non-negative matrix factorization is being used to get this *X*. Here, *X* and *X*^*T*^ are considered equivalent to each other. We pick *X* here to calculate the quantification strategy for it further.

#### 3.13.2 Quantification Strategy

In this method, as each row of *X* defines the probability distribution of the corresponding nodes, we can find the most uncertain nodes based on the values of *X*. We have used the information entropy and the Jensen-Shanon divergence of the node probability distribution to obtain the quantification measure and formulate the significance of edges. These two measures can be defined as the following:

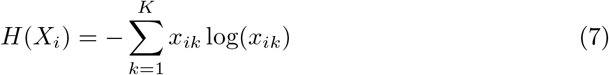

Finally, the edge weight between a pair of nodes is defined as ℒ ℰ _*ij*_ and represented below:

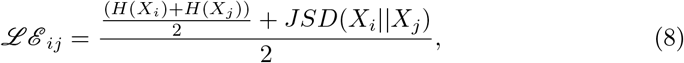

where *H*(*X*_*i*_) is the information entropy of node *i, X*_*i*_ is the probability distribution of node *i* and *JSD*(*X*_*i*_|| *X*_*j*_) is the Jensen-Shanon divergence between *X*_*i*_ and *X*_*j*_. Thus, the whole undirected (initially assumed) heterogeneous network is weighted by this method. We only assign these link entropy weights to the unweighted edges. The other two previously assigned weights (i.e., for pathway-pathway and gene coexpression networks) are the same as earlier.

### 3.14 Identifying all different entities and build word2vec embedding of all the unique entities BioBERT model

Different entities are used to construct these disease-specific heterogeneous networks. We first collectively identify these other kinds of unique entities, and their concepts are formed by appending additional information. These various types of entities are, namely, pathway, only hyper/hypo methylated genes, only up/downregulated genes, down/up-regulated hypermethylated genes, down/up-regulated hypomethylated genes, SNP variants, and all amyloid proteins, mapped to their UniProt identifiers, PTMs, and ICD-CM (i.e., E11.8 and G20) codes. There are a total of 31918 unique entities present in the T2D network and 47561 unique entities present in the PD network. We have used the BioBERT (Bidirectional Encoder Representations from Transformers for Biomedical Text Mining) [50] model to convert entity-associated concepts to vector embeddings. So, finally, we get an initial vector embedding for each unique entity for the individual heterogeneous networks.

### 3.15 Applying Heterogeneous Graph Transformer Network (HGT) to individual disease-specific heterogeneous networks to perform link analysis

After building the individual disease-specific networks for T2D and PD, we infer the plausible links between DMGs and PTMs for each network respectively. As PTM is one of the most critical factors causing protein aggregation, unknown links associated with PTMs are crucial for our investigation. These findings are due to the participation of protein aggregation, which results in both T2D and PD in patients. Likewise, DNA methylation is another complex mechanism that plays a crucial role in causing the same co-morbid diseases. Hence, to figure out the plausible latent cause of these two co-morbid diseases, link analysis between DMGs and PTMs is essential. We use the graph neural network-based method, HGT (Heterogeneous Graph Attention Network), for our proposed individual heterogeneous network associated with T2D and PD. The motivation for using this model for our network analysis is its potential for link prediction while capturing heterogeneous attribute information in the network. This HGT method was first introduced by Hu and his group in their paper [33] for node classification and link analysis tasks in a heterogeneous graph. The heterogeneous graph consists of different types of nodes and edges. Hence, this type of network always contains rich semantic information. So, the mere application of a simple graph-based deep learning approach is not suitable for learning these networks.

We define the HGT model by our disease-specific heterogeneous graph 𝒢, which is made up of 𝒱, ℰ, 𝒜, ℛ. Each node *v* ∈’ 𝒱, and each edge *e*∈’ ℰ has a mapping function *f* (*v*) that goes from 𝒱, to 𝒜 and *ϕ*(*e*) that goes from ℰ to ℛ. Here, *V* denotes the entities, and *E* denotes the biological relations in the heterogeneous graph *G*. An edge *e* = (*s, t*) linked from source node *s* to target node *t*, whose meta-relation is denoted by *< f* (*s*), *ϕ*(*e*), *f* (*t*) *>*, and the inverse relation is referred to as *ϕ*(*e*)^−1^. Specifically, the node represents the genes (including methylated ones), pathways, SNPs, PTMs, and amyloid proteins (including central amyloids). The edges represent the interrelations among the connected nodes. So, the meta-relations are different for each type-specific edge. For instance, *pathway-methylated* refers to the connection between the relation between node pairs *pathway* and *methylated gene*. The HGT extracts all the biologically linked node pairs of target node *t* and source node *s* via the edge *e*. The output of the *l*− *th* layer is denoted as *H*^*l*^, fed to the (*l* + 1) −*th* layer as input. The end node representations of the whole graph *H*^*L*^ can be obtained while stacking the *L* layers. The input heterogeneous graph with the transformer architecture is shown in **Figure 2**. The mutual attention between the node pair *s* and *t* is calculated through the following GNN attention mechanism:

**Fig. 2.**
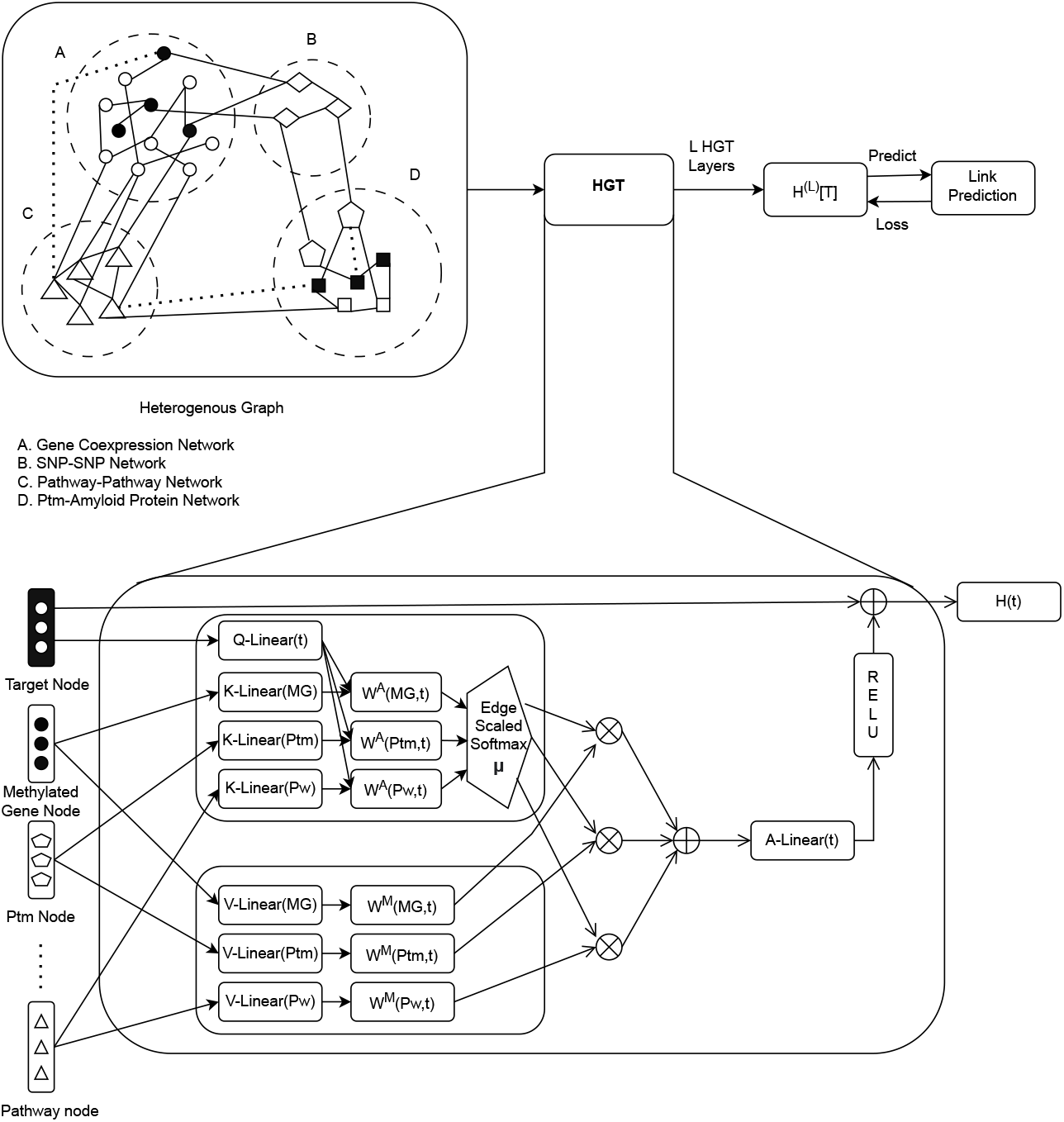
Proposed heterogeneous transformer architecture

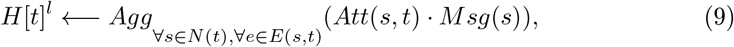

where *Att* estimates the attention of the source node using an additive mechanism, *Msg* refers to the information propagated through only the source node, and *Agg* aggregates the neighborhood message by the attention weight that follows the nonlinear activation function. To calculate the attention, we measure the dot product of the target node *t* and the source node *s* after mapping *t* to a query vector *Q* and *s* to a key vector *K*. Due to the heterogeneous nature of the graph for different biological relation, we calculate the *h* − *head* attention as follows:

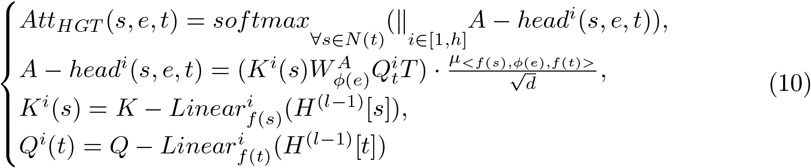

For the *i* − *th* attention head *A* − *head*^*i*^(*s, e, t*) we project the *f* (*s*) type node *s* into the *i* − *th* vector 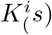 applying a linear projection 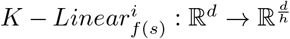 to move the *i*− *th* attention head *A*− *head*^*i*^(*s, e, t*) into the *i*− *th* key vector *K*^*i*^(*s*). Here, *h* is the number of heads, and 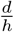 is the number of dimensions of each head. As in our disease-specific heterogeneous graph, each node type has a different linear projection to reflect the distribution differences throughout the model. Hence, 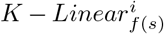is indexed by the source node’s type *f* (*s*) and 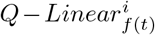 linear projection is indexed for *i* − *th* target node. For different edge type-specific matrix 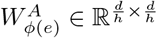 for each of the edge types *ϕ*(*e*), as the attention weight to measure the similarity between query vector *Q*^*i*^(*t*) and key vector *K*^*i*^(*t*) instead of using simple dot product between these two vectors. For instance, in **Figure** 2 the DNA methylated gene edge attention matrix 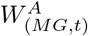, for PTM 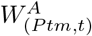, and for pathway 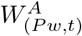 are shown. For each node pair (*s, t*), multi-head message passing is employed as follows:

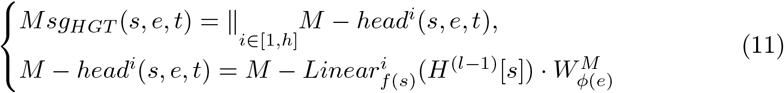

Hence, to obtain the *i*−*th* message head *M* −*head*^*i*^(*s, e, t*), we project *f* (*s*) message vector with 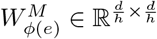 matrix to capture the edge dependency. To obtain each of the target node *t*’s attention vector, softmax is employed to sum up altogether to one and used to average the messages coming from the source node that leads to producing updated vector Ĥ ^*l*^[*t*] as below:

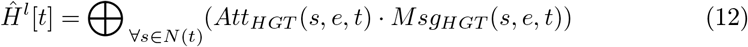

This aggregation fetches all the different types of source node information and aggregates them to the target node t. In the final step, for each target node, *t* is mapped to each node type *f* (*t*). This is calculated by applying *A*−*Linear*_*f*(*t*)_ projection to the updated vector Ĥ ^*l*^[*t*] with the non-linear activation on it and the residual connection as well:

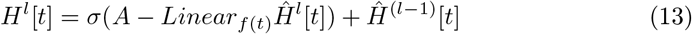

Finally, the *H*^*L*^ representation is obtained by stacking HGT blocks for *L* layers, and that is used to perform the link prediction between the desired node pairs for each disease-specific heterogeneous network.

## 4 Experiments

To initiate the experiment, at first, the entire dataset with their corresponding entity embeddings for T2D and PD individually are converted into DGL API [76] compatible data format. Then, each dataset is split into train, valid, and test sets. Next, the Open-HGNN [27] framework is used for our experiment. We use the Heterogeneous Graph Transformer Network (HGT) graph neural network model to perform the link prediction tasks for individual disease-specific heterogeneous networks. Then, we generate the desired target link-specific comparative ROC-AUC (Receiver Operating Characteristic Area Under Curve) accuracy scores for each network. We also predict some novel common pathway signatures relating to T2D and PD in the experiment. We further validate these findings with the available literature.

### 4.1 Link Prediction for Desired Target Relations

Our constructed individual disease-specific weighted heterogeneous networks associated with T2D and PD are used independently for link analysis tasks, to infer the plausible common target links between these two prevalent diseases. The edge weights calculate the message passing through the graph neural network. To perform the link analysis using the HGT model, the first task is to form the meta-paths between the different entities in the network. The meta-paths for T2D are comprised of the nodes namely, pathway, gene, downregulated_ hypomethylated_ gene, upregulated_ hypomethylated_ gene, hypomethylated_ gene, snp, amyloid_ protein, ptm, and central_ amyloid_ protein_ with_ ICD-E11.8. Similarly, for PD the meta-paths contain pathway, gene, downregulated_ gene, upregulated_ gene, downregulated_ hypomethylated_ gene, upregulated_ hypomethylated_ gene, hypomethylated_ gene, snp, amyloid_ protein, ptm, and central_ amyloid_ protein_ with_ ICD-G20. The elaborate description of individual relations with corresponding meta-path ID between entity pairs of the diseases T2D and PD are given in the Supplementary file.

In each of the specific disease-based heterogeneous networks, a desired target link is considered and link analysis is performed using the HGT model individually for both diseases. We use the OpenHGNN [27] framework for implementing our T2D and PD-specific heterogeneous network-based HGT model. In the case of T2D, we split the whole constructed dataset as 70% for training, 10% for validation, and the remaining 20% for testing. The dataset associated with PD is being split as 60% for training, 20% for validation, and the remaining 20% for testing. To get the performance of each relation-specific link for each disease, roc_auc accuracy is calculated by optimizing the parameters using the validation dataset. For the HGT model, we take the hidden dimension as 128, the number of heads as 8, the learning rate as 0.005, and weight decay as 0.001. In association with that, the Adam optimizer is used for tuning the parameters.

## 5 Results

To our knowledge, no computation method exists for finding hidden co-morbid patterns from heterogeneous multi-omics networks associated with T2D and PD. However, we perform our method’s comparative analysis with the Heterogeneous Graph Attention Network (HAN) [77] model. From the result in Tables 1 and ‘2, we can see visibly good accuracy scores due to the higher number of link instances in the individual networks. For example, the target link *pathway-central_amyloid_protein_T2D_E11*.*8* for T2D gives a good result of 0.89 accuracy. Similarly, the relation *pathwaycentral_amyloid_protein_pd_G20* also provides a good accuracy score of 0.85. Our proposed methodology infers the hidden biological connections between T2D and PD based on common pathway signatures. T2D and PD module-specific pathway sub-networks are merged to obtain the final common signature. Thus, we get the overlapped known pathway links to exit among these two diseases. The novel hidden common signature analysis is done on two factors, namely hypomethylation and PTMs linked to familiar predicted pathway entities individually for T2D and PD. The new findings of common pathway signatures for hypomethylation, PTMs, and, finally, in association with the known pathway links are given in the subsequent sections below.

**Table 1:** Target relation-specific roc_auc scores for T2D.

| Target<br>Links for T2D | Models |  |
| --- | --- | --- |
|  | HGT<br>(roc_auc) | HAN<br>(roc_auc) |
| pathway-hypo | **0.83** | 0.75 |
| pathway-central_amyloid_protein_T2D_E11.8 | **0.89** | 0.80 |
| central_amyloid_protein_T2D_E11.8-ptm | **0.87** | 0.78 |

**Table 2:**
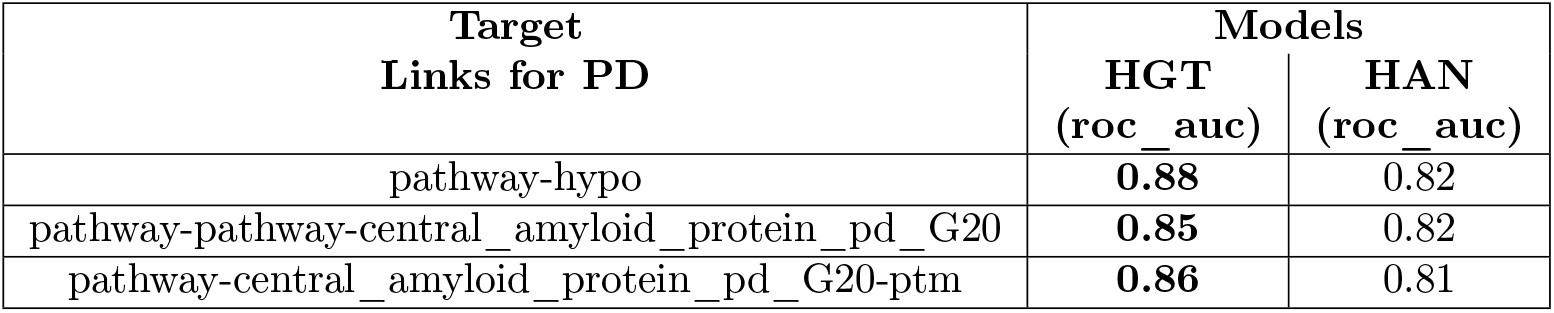
Target relation-specific roc_auc scores for PD.

| Target<br>Links for PD | Models |  |
| --- | --- | --- |
|  | HGT<br>(roc_auc) | HAN<br>(roc_auc) |
| pathway-hypo | **0.88** | 0.82 |
| pathway-pathway-central_amyloid_protein_pd_G20 | **0.85** | 0.82 |
| pathway-central_amyloid_protein_pd_G20-ptm | **0.86** | 0.81 |

### 5.1 Newly predicted common pathway signature between hypomethylated genes for T2D and PD

Our proposed method predicts the common pathway signatures linked to the hypomethylated genes for T2D and PD. Among all the newly predicted pathway signatures for both the co-morbid diseases, few have been found in the literature (see the supplementary Table S3 and S4, available online). To cite an example, a predicted instance is shown in the supplementary Figure S2 (available online). It is observed that in Figure S2(a), the common pathway pattern *REAC-TOME_INNATE_IMMUNE_SYSTEM* is linked with the hypomethylated genes VIPR2/VPAC2 for T2D and QRSL1 for PD. To further clarify the finding we search for its basis in the existing literature. It is already a known fact that vasoactive intestinal peptide (VIP) can stimulate glucose-dependent insulin secretion by binding to VIPR2 receptors [31]. In another literature [85], it is shown that pituitary adenylate cyclase-activating peptide (PACAP) can activate VPAC1 and VPAC2 (i.e. VIPR2) receptors and in association with that, the VPAC1-mediated hepatic glucose increase prevents the increased insulin secretion by VPAC2 activation. So, PACAP derivatives such as VPAC2 can efficiently promote glucose-dependent insulin secretion and safeguard islet beta cells. It legitimately defines VIPR2 as an important target gene that can be associated with T2D. Now, the hypomethylation of genes is already proven to be related to DNA damage and oxidative stress, leading to beta cell dysfunction [74]. This hypomethylation can be observed at target genes and can be explained through competing for proapoptotic and anti-apoptotic processes during ER (Endoplasmic Reticulum) stress response in diabetic islets. The researchers further report that specifically an altered DNA methylation profile in the pancreatic islets of T2D patients with a major dominance of hypomethylation in sequences outside CpG islands (CGIs). Hence the dysregulation of genes may notably contribute to beta cell functionality, cell death, and adaptation to metabolic stress. This concludes that hypomethylation of VIPR2 can significantly affect the performance of beta cells in T2D patients. In connection regarding the gene QRSL1 and its hypomethylation related to PD can similarly be verified by few literature [21, 22]. Mitochondrial dysfunction plays a crucial role in causing PD, but the underlying mechanism still needs to be addressed. Various facts show that QRSL1 is a rare, damaging gene variant accountable for mitochondrial dysfunction leading to PD in patients. In light of keen observation, it may be inferred that the genomic global hypomethylation of QRSL1 may play a key role in the causation of mitochondrial dysfunction. The activation of the innate immune system is closely related to the pathogenesis of T2D [59]. This study analyses the complex interplay between inflammation and neuronal dysfunction associated with T2D. Neuronal death can induce inflammation while releasing apoptotic and necrotic cellular factors. These factors can be recognized by innate immune cells [38]. The activation of innate immune cells by different verified factors can then introduce an adaptive immune response which leads to selective and specific patterns of neuronal injury. This neuronal dysfunction carries an important role in the pathogenesis of PD. In a few works, [6, 66] show that the innate immunity system pathway is related to VIPR2 and QRSL1. So, this biological literature verifies our predicted links of innate immunity system pathway to hypomethylated genes in the relevance to link the co-morbid signature between T2D and PD.

### 5.2 Newly predicted common pathway signature between PTMs for T2D and PD

Few of our predicted common pathway signature links to PTMs show a literature validation for predicted PTM links association to T2D and PD. For instance, in Figure S2(b), the *REACTOME_EGFR_DOWNREGULATION* is linked to two PTMs, namely *phosphorylation* and *acetylation*. It is observed that [71], the primary client protein of the beta cell ER is proinsulin. To promote its perfect folding, the consequence of entering the proinsulin in ER is bound reversibly by molecular chaperones, namely, BiP. When free BiP levels fall due to its binding to newly produced proteins, this triggers the PERK gene to phosphorylate eIF2*α*, and protein translation fails during ER stress. In the scenario of PERK, reduced protein synthesis prevents new proteins from entering the ER when nothing can be correctly folded. Due to the defect of insulin folding, T2D can occur. Another incident occurred due to the activation of the insulin receptor signal in the cell’s interior via the phosphorylation of target molecules, including insulin receptor substrate 1 (IRS1) on tyrosine residues. This can be blocked if phosphorylation occurs in IRS1 on serine residue by the Jun-N-terminal kinase (JNK) and triggers in the peripheral tissue of obese individuals. So, phosphorylation participates in the formation of protein misfolding, leading to T2D. One recent work proposed by Yakhine et al. [80] is significant in showing the relationship between protein acetylation and PD. According to the authors, acetylation is a post-translational modification regulated by antagonistic enzymes, histone acetyl-transferases (HATs), and histone deacetylases (HDACs). HATs transfer the acetyl group from acetyl-COA to lysine residues of proteins, whereas HDACs remove it. The impairment of HAT and HDAC activities causes changes in cellular processes, which leads to PD in patients. Hence, the acetylation level of proteins leads to cell death in idiopathic Parkinson’s disease individuals. The association between the EGFR pathway and the acetylation is observed in the work by Song et al. [67]. As alterations of epidermal growth factor receptors are involved in various human diseases, the EGFR pathway plays an essential role in developing T2D and PD. Another study by Wee et al. [78] shows that adding EGF Hela cells activates the EGFR to cause global phosphorylation. The connection between the EGFR pathway and protein acetylation is observed in the study by Kim et al. [42]. This verifies that our proposed prediction for the EGFR pathway and acetylation leads to PD in healthy individuals.

### 5.3 Common pathway signature linked to the newly predicted pathway signatures for T2D and PD

To show the final common pathway signature connected to the predicted pathway signatures of T2D and PD, we retrieve the commonly known connection from the common pathway-pathway interactions. Among all the connected signatures, we obtained a few findings verified with justified biological validations. For instance, in Figure S2(c) the known common pathway *KEGG_GLYCOLYSIS_GLUCONEOGENESIS* is connected with the predicted pathway signatures namely, *REACTOME_INNATE_IMMUNE_SYSTEM* and *REACTOME_EGFR_DOWNREGULATION*. One report [63] shows that differential glycogen and glucose metabolism in dendritic cells within the innate immune response, concluding to their interrelated pathway connections. Catabolic pathways, including glycolysis and fatty acid oxidation (FAO), are interconnected with biosynthetic and redox pathways. Innate immune cell activation and differentiation trigger immense metabolic changes to support their functional activities [54]. In some studies, it has been noticed that increased glycolytic activity authenticates the formation of many complex diseases. Pyruvate dehydrogenase (PDH) complex acts as a gatekeeper between glycolysis and oxidative phosphorylation, and activation of PDH inhibits glycolytic activity. Another study by Jeon et al. [35] shows that EGFR signaling pathways play a crucial role in the upregulation of large-sized glycosomes in diseased cells, which functionally govern the promotion of glycolysis-derived biosynthesis. These validations further show the plausible connection of hidden co-morbid factors for T2D in association with PD in patients.

## 6 Discussion and Conclusion

This paper proposes a novel approach for building disease-specific heterogeneous networks while integrating different tissue-specific information and identifying the common co-morbid patterns between T2D and PD. Here, we choose common pathway signatures based on hypomethylation and PTM as these co-morbid patterns. To the best of our knowledge, the proposed approach is the first method of its kind, where we build two different tissue-specific heterogeneous networks by integrating transcriptomics, epigenetics, and epistasis data for both of these co-morbid diseases. We refer to these tissue-specific sub-networks as local networks. Then, we connect these two heterogeneous networks with the Amyloid-PTM interactome (i.e., PTM-AmyNet) and refer to it as a global network. Our method shows various advantages in the view of heterogeneous data fusion and how these heterogeneous networks help to infer new latent co-morbid patterns between T2D and PD. First, we perform the tissue-specific microarray-based gene-expression analysis and build a set of weighted gene modules or gene expression subnetworks using WGCNA. Then, based on those modules, we build weighted pathway-pathway sub-networks for each module. In the pathway-pathway sub-networks, the inter-related edge weights are calculated using the mutual phenotypic information of the gene-expression specific values and semantic similarity. To reflect the optimal pathway importance in the corresponding edge weights, we take the maximum of the phenotypic score and their mean scores and sum it up with the semantic score to get the final weight of individual pathway pairs. This technique incorporates the importance of the pathway in a specific relation associated with the similarity value of gene ontology scores. Second, to obtain the overlapping genes with differentially methylated genes, we perform the DNA methylation profiling of the same tissue-specific gene expression microarrays individually for T2D and PD. Third, we build the epistasis network using the GWAS-based query SNPs for mapped traits such as T2D and PD for the desired population. These query SNPs are further used to fetch proxy SNPs with the MAF scores ≥ 0.05. The final SNP-SNP sub-network is formed using the PLINK tool by considering the MAF scores for binomial probabilistic random genetic distribution to build the sample genetic weight matrix for PLINK analysis. As the MAF scores with ≥ 0.05 are already verified to be responsible for complex diseases like T2D and PD, we select them as the probabilistic score for creating the genetic weight matrix. Hence, these probabilistic values are reflected in the selection of SNP variants. These SNP-SNP network variants are further analyzed with the Ensembl-VEP tool to obtain disease associations and genetic links. After building this heterogeneous network, we create a unique entity concept for each node entity. Then, BioBERT embedding is used to get the context-specific unique embedding for each entity ID. This approach helps to give the model type-specific information for the corresponding nodes in the individual heterogeneous networks. We try to capture the topological information of the network based on the importance of the entities in the network. So, we calculate the link entropy of the whole network to obtain the link weight of all the inter- and intra-related relationships in the network. Only we keep the gene-gene and pathway-pathway edge weights the same as we calculated earlier, and the remaining edges are weighted according to the obtained link entropy weights. This method incorporates the entity-specific information according to the gene expression values and the topological significance of the remaining sub-networks. Besides building tissue-specific individual disease-based heterogeneous networks by integrating multiple genomic level information through a unique approach, our method also further bridges the latent factors, namely, hypomethylation and PTM based on unique pathway signatures. Few of these predictions have also been biologically validated, and the remaining findings may need more important biological investigations.

Despite the novel co-morbid pattern predictions from our novel method of building disease-specific heterogeneous networks, there is still room for improvement in the work. In our method, due to the number of target relation-specific instances for individual diseases, our proposed network may lack some vital information that may lead to a few more exciting outcomes. We may further integrate more of these instances to infer more unknown and valuable results in the future. Our proposed approach uses the HGT graph, a neural network model, to learn the individual heterogeneous networks and obtain link prediction scores of target relationships. We may further train our network with other heterogeneous graph neural network models for a comparative analysis of individual relationships. Another modification can be made to the negative sub-graph construction methodology to get more critical latent patterns from the network. Finally, we hope that our proposed work will be helpful in the integration of tissue-specific transcriptomics, epigenetics, and epistasis-related data in association with the amyloid-PTM network for T2D and PD and infer hidden common co-morbid patterns that can be targeted as potential biomarkers for a better prognosis of these prevalent diseases. However, our findings may require further biological intervention for better verification. These latent pattern predictions may be further helpful in contributing to the research of drug therapeutics to treat the target factor responsible for creating T2D and its co-morbid condition, PD, at the same time in patients.

## Data Availability

All data produced in the present study are available upon reasonable request to the authors.

## Supplementary information

The Supplementary file is provided along with the manuscript.

